# Genetic analyses of blood pressure traits in 140,000 admixed adults from Mexico City

**DOI:** 10.64898/2026.09.17.26363296

**Authors:** Michael Turner, Jaime Berumen, Sam Morris, Diego Aguilar-Ramírez, Luisa Fernández-Chirino, Tin Orešković, Paulina Baca, Elizabeth Barrera, Fernando Rivas, Louisa Gnatiuc Friedrichs, Michael Hill, Eirini Trichia, Alejandra Vergara-Lope, Rachel Wade, Ernst Mayerhofer, Moeen Riaz, Iona Y. Millwood, Robin G. Walters, Alfred Pozarickij, Jerome I. Rotter, Jie Yao, Henry J. Lin, Xiuqing Guo, Natalie Staplin, Pablo Kuri-Morales, Richard Haynes, Jesús Alegre-Díaz, Jonathan R. Emberson, Roberto Tapia-Conyer, Jason M. Torres

## Abstract

Blood pressure (BP) is a major, heritable determinant of cardiometabolic health. Genome-wide association studies (GWAS) have identified thousands of BP-influencing genetic variants. People of admixed American (AMR) ancestry harbour variants that are rare in other populations yet remain understudied in GWAS. Here, we perform GWAS of four BP traits in 140,559 participants from the Mexico City Prospective Study. Conditional analyses identified 78 independent signals for systolic BP, 61 for diastolic BP, 88 for mean arterial pressure and 24 for pulse pressure. Trans-ancestry meta-analysis externally replicated 13 novel signals, and six more replicated in AMR-ancestry meta-analysis. Overall, we found significant heterogeneity between AMR and other ancestries at nine loci. Functional integration analyses implicated the heart, arterial tree, adrenal glands, and kidney in BP regulation. Analysis of exome-sequenced rare variants identified genes that may act through kidney and endothelial function. The results underscore the value of including diverse populations in genetic discovery.

## Main

Elevated blood pressure (BP) is heritable^1^ and associated with cardiovascular and other diseases^2–6^. Together such diseases cause an estimated 11 million deaths annually^7^. Lowering BP reduces morbidity and mortality^8^ and BP-lowering interventions are made possible by a detailed physiological understanding of blood pressure regulation.

A genome-wide association study (GWAS) in over one million people of European ancestry identified 2103 independent signals explaining over 60% of single nucleotide polymorphism-based heritability of BP traits^9^. GWAS with other ancestries frequently corroborate variant associations seen in European-ancestry populations and also identify novel BP-influencing variants^10–12^. These, and other^13–15^, studies have demonstrated the complex, polygenic architecture of BP and indicated roles for vascular, kidney and neuro-endocrine tissues in BP regulation.

Latin America accounts for roughly 8% of the global population^16^. Complex patterns of admixture between peoples of African (AFR), European (EUR) and Indigenous American (IAM) ancestries have led to allele frequencies and patterns of linkage disequilibrium (LD) that differ from those in other populations^17,18^. However, individuals of Latin American descent (often referred to as “Hispanic” or “Latino”) who have admixed American (AMR) genetic ancestry – are among the least represented in genetic research^16^. Despite their low representation in GWAS, AMR-ancestry populations have made important contributions to the understanding of complex traits that influence health^12^. For example, variants in *SLC16A11* that increase risks for type 2 diabetes are far more common in AMR-ancestry populations than in others^19^.

Here, we perform GWAS of systolic BP (SBP), diastolic BP (DBP), mean arterial pressure (MAP) and pulse pressure (PP) in roughly 140,000 participants from the Mexico City Prospective Study (MCPS)^20,21^. This represents the largest effective sample size to date for an AMR-ancestry GWAS of blood pressure. Our sample size surpasses the numbers of AMR-ancestry participants in the Million Veteran Program (58,000)^22^ and the All of US study (42,000)^23^. We perform conditional analyses to identify independent signals and use approximate Bayesian fine-mapping techniques to infer causal variants. We further replicate novel signals in external cohorts from both AMR and non-AMR ancestry populations and evaluate the tissues implicated by BP-associated variants using several functional integration methods. Finally, we explore the influence of rare coding variants on blood pressure traits through exome-wide association analyses and gene-based tests.

## Results

### Genome-wide association studies

GWAS of 42,121,162 well-imputed variants (imputation r^2^ ≥0.4 and N_eff_ ≥30, see Methods) in 140,559 individuals with plausible BP measures were conducted with SUGEN^24^ to account for the extensive relatedness and complex admixture present in MCPS^18^. Participant characteristics are summarised in Supplementary Table 1.

Genomic inflation was well controlled, as shown by covariate-adjusted linkage disequilibrium score regression^25^ (cov-LDSC) intercepts of 1.032, 1.031, 1.037 and 0.995, for SBP, DBP, MAP and PP, respectively (Supplementary Fig. 1). Further, cov-LDSC demonstrated heritability estimates of 11.3% for SBP and 9.9%, 11.4% and 6.3% for DBP, MAP and PP, respectively. These are lower than estimates from European-ancestry GWAS, but in line with the lower heritability estimates previously seen in AMR-ancestry populations^26^.

For SBP, GWAS significant (p < 5×10^-8^) associations were found at 72 independent loci (genetic regions around GWAS-significant variants at least 500kb from other significant variants), with 62 loci known and 10 novel (Fig. 1A, Supplementary Table 2). Stepwise conditional analyses identified 78 independent signals at these loci, with 14 having previously-reported lead variants (see Methods for full details). In addition to the 10 signals at novel loci, 9 signals at known loci were conditionally-independent of all known variants previously associated with any BP trait (LD r^2^ <0.1, p-value_conditional_ <5×10^-8^, Supplementary Table 3). These 9 signals were considered novel and taken forwards for external replication alongside those at novel loci.

**Figure 1.**
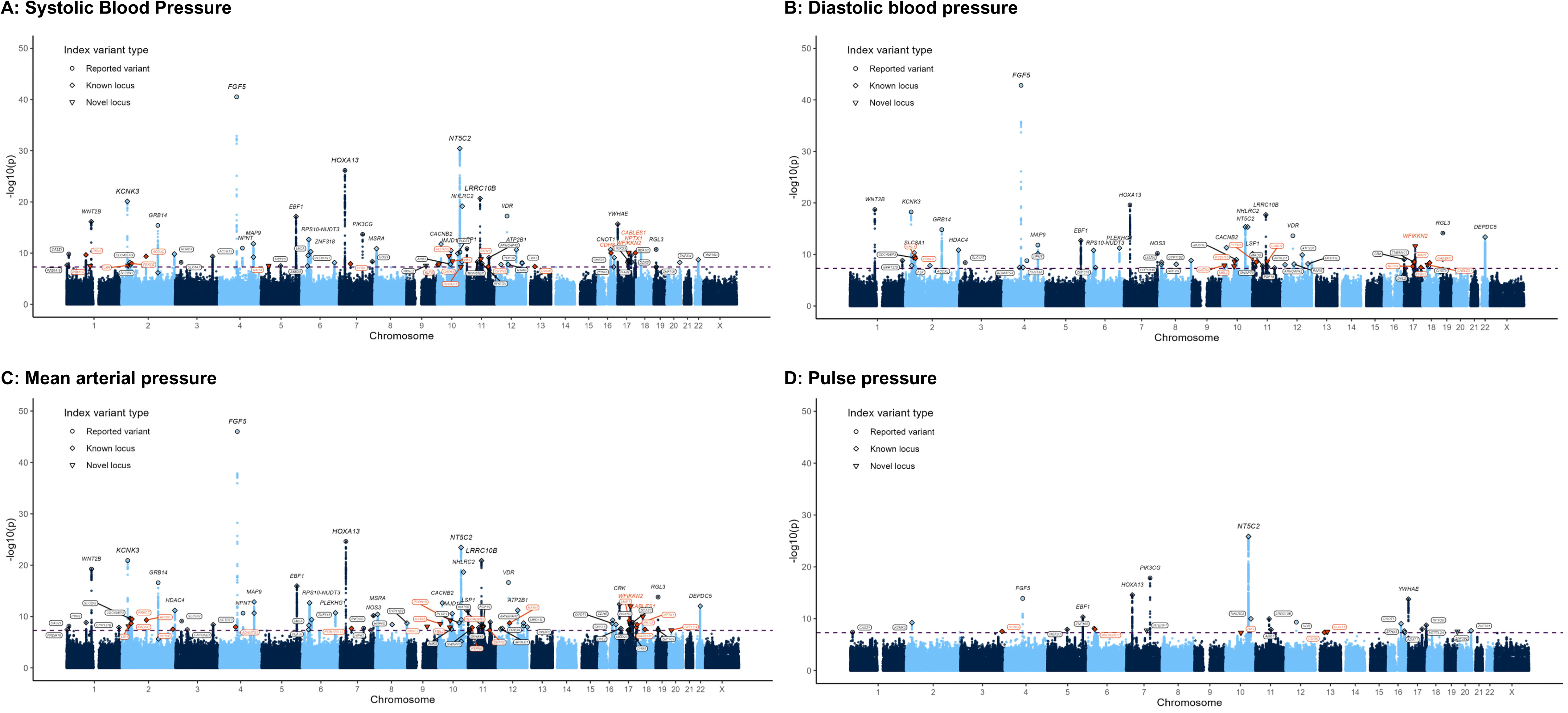
Manhattan plots of genome-wide association studies of blood pressure traits. Panels show genomic position across the 23 chromosomes on the X axis and -log_10_(p-value) on the y axis for genome-wide association studies of systolic blood pressure (A), diastolic blood pressure (B), mean arterial pressure (C) and pulse pressure (D). Lead variants are labelled with the name of the nearest canonical gene and surrounded by a circle if they are a previously-reported variant, a diamond if they are at a known locus and by a triangle if they are at a potentially-novel locus. Shapes and gene names are orange if they have ≥2 supportive associations within 500kb and are either at a novel locus or are conditionally-independent of all known associations at a known locus. Shapes and gene names are black if these criteria are not met.

For DBP, significant associations were found at 60 loci, including 6 novel loci (Fig. 1B). Conditional analyses identified a single secondary signal in *GRB14* (Supplementary Table 4), with 7 signals conditionally independent and considered novel (Supplementary Table 5). GWAS of MAP (Fig. 1C) and PP (Fig. 1D) identified significant associations at 82 loci (9 novel) and 28 loci (2 novel), respectively. For MAP, conditional analyses identified 88 signals at these 82 loci (Supplementary Table 6), including 3 independent signals near *NT5C2* (and pairs of independent signals at four other loci), and 13 signals conditionally independent and considered novel (Supplementary Table 7). No secondary signals were identified for PP (Supplementary Table 8), but 3 signals were conditionally independent and considered novel (Supplementary Table 9).

As expected given their inter-related nature, there was substantial overlap between significant associations for the four BP traits examined with 114 distinct signals identified (Fig. 2, Supplementary Table 10). DBP and MAP were most closely aligned, with 55 of 61 DBP signals (90.2%) also identified as MAP signals and cov-LDSC genetic correlation of 0.994 (Supplementary Table 11). Similarly, 65 of 78 SBP signals [83.3%] corresponded to MAP signals (genetic correlation 0.991), and 21 of 24 PP signals [87.5%] corresponded to SBP signals (genetic correlation 0.934). In contrast, PP was especially poorly correlated with DBP, with only 9 of 24 PP signals (37.5%) corresponding to DBP signals (genetic correlation 0.813).

**Figure 2:**
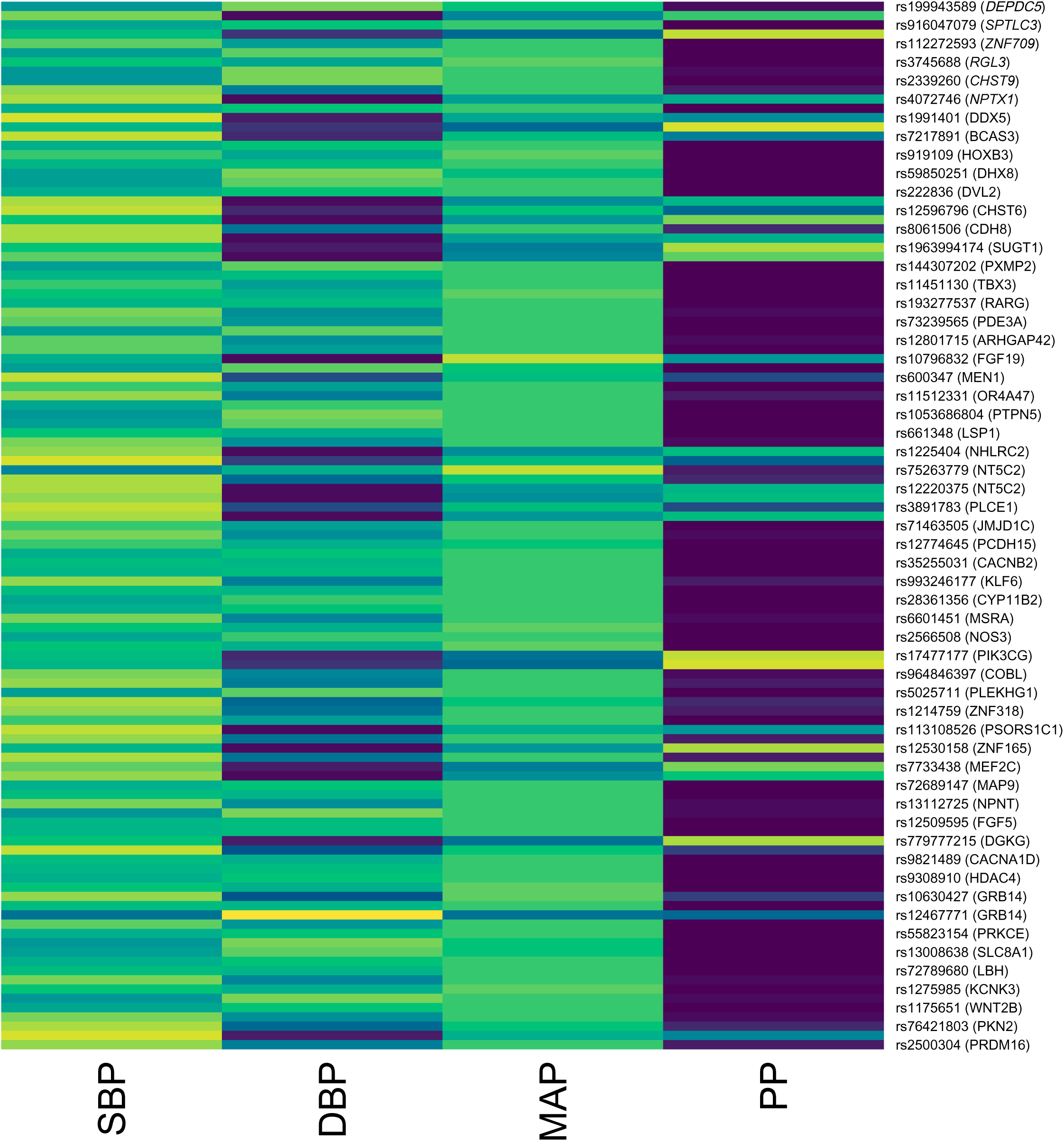
Strength of association across 114 distinct signals for four blood pressure traits. Image displays a rectangle for each blood pressure trait at each signal. The colour corresponds to the strength of the association with brighter, with darker colours corresponding to stronger associations. For alternate signals rsIDs and gene names are displayed on the right. These data are also tabulated in supplementary table 10. DBP: diastolic blood pressure, MAP: mean arterial pressure, rsID: reference single nucleotide cluster identifier, PP: pulse pressure, SBP: systolic blood pressure.

In keeping with previous studies, there was a clear relationship between allele frequency and effect size with rarer GWAS-significant variants having larger effect sizes (Supplementary Fig. 2)^27^. Cumulatively, common (minor allele frequency [MAF]>5%) significantly-associated variants explained 1.8%, 1.4%, 2.0% and 0.52% of the variance observed in MCPS for SBP, DBP, MAP and PP, respectively. Low-frequency (0.1%≤MAF≤5%) and rare variants (MAF<0.1%) together explained 0.15%, 0.10%, 0.17% and 0.16% of the variance in SBP, DBP, MAP and PP, respectively. These results are consistent with previous work showing that common variants explain most of the phenotypic variance in BP traits^9^. The vast majority of associated variants were not in protein-coding regions (Supplementary Table 2, Supplementary Tables 4, 6 and 8). Specifically, only the previously reported variant in *RGL3* (chr19:11416089:T:G/rs167479, predicted to cause a missense histidine to proline substitution) associated with SBP, DBP and MAP had a lead variant in a protein-coding region.

### Previously-reported variants showed no ancestral heterogeneity

To assess heterogeneity of genetic effects between ancestries, associations of previously-reported variants were obtained from GWAS studies of non-AMR individuals in the GWAS catalog. Of the 14 lead variants for SBP signals that were previously-reported, 11 had associations described in sufficient detail for heterogeneity assessment^28^ and none showed significant heterogeneity (all p_het_ >0.1, Supplementary Fig. 3, panel a). Similarly, there was no significant heterogeneity for DBP, MAP and PP (all p_het_ >0.1, Supplementary Fig. 3, panels b-d).

### Fine mapping

To identify the most likely causal variant at each signal, we fine-mapped genetic associations using an approximate Bayesian approach^28^ (see Methods for full details). For SBP, 12 of the 78 (16%) signals had a maximum posterior probability of association (PPA) of >0.5, and 4 of these resolved to a single likely causal variant (Supplementary Table 12). These signals have all been reported previously and include the signal at the *FGF5* locus, which has the strongest association in MCPS, and the signal at the *VDR* locus that was first reported in 2023^29^.

For DBP, 11 of 61 signals (19%) had a maximum PPA >0.5 (with 4 resolving to a single variant, Supplementary Table 13). For MAP, 17 of 84 signals (20%) had a maximum PPA >0.5 (4 resolving to a single variant, Supplementary Table 14). For PP, 5 of 23 signals (21%) had a maximum PPA >0.5 (1 resolving to a single variant, Supplementary Table 15).

### Replication of novel signals

Replication was conducted through inverse variance-weighted meta-analysis of summary results from cohorts with AMR-ancestry participants (the Multi-Ethnic Study of Atherosclerosis [MESA], Million Veteran Program [MVP], and pan-UK Biobank [pan-UKB]) and cohorts composed exclusively of other ancestries (Biobank Japan [BBJ], China-Kadoorie Biobank [CKB]).

Of 19 novel SBP signals (10 at novel loci and nine at known loci independent of all reported variants), two (rs964846397, rs993246177) were not present in any other cohort. Both are rare variants present only in IAM local ancestry segments in MCPS (Supplementary Table 16 and MCPS variant browser [URLs]). Of the remaining 17 signals, four met replication criteria (directionally consistent and false discovery rate [FDR]<5%, Table 1, Supplementary Table 17). These included rs600347 upstream of *MEN1* (Fig. 3A), which is more common Latin America than Europe and Africa (Fig 3C) and in IAM-ancestry genetic segments in MCPS (Fig. 3D). The other three were rs55823154 in an intron of *PRKCE* (Extended Data Fig. 1A), rs1561477 in an intron of *ACOXL* (Extended Data Fig. 1B), and the intergenic variant rs903845 near *RAI14* (Extended Data Fig. C). Notably, the signal at *PRKCE* was significantly stronger in external AMR-ancestry cohorts, with an effect size of -0.30 mmHg (95%CI -0.44 to -0.16) per G allele in the AMR-ancestry replication meta-analysis vs -0.09 mmHg (-0.14 to -0.04) in the non-AMR-ancestry replication meta-analysis (p_het_=0.005, Extended Data Fig. 1A).

**Figure 3:**
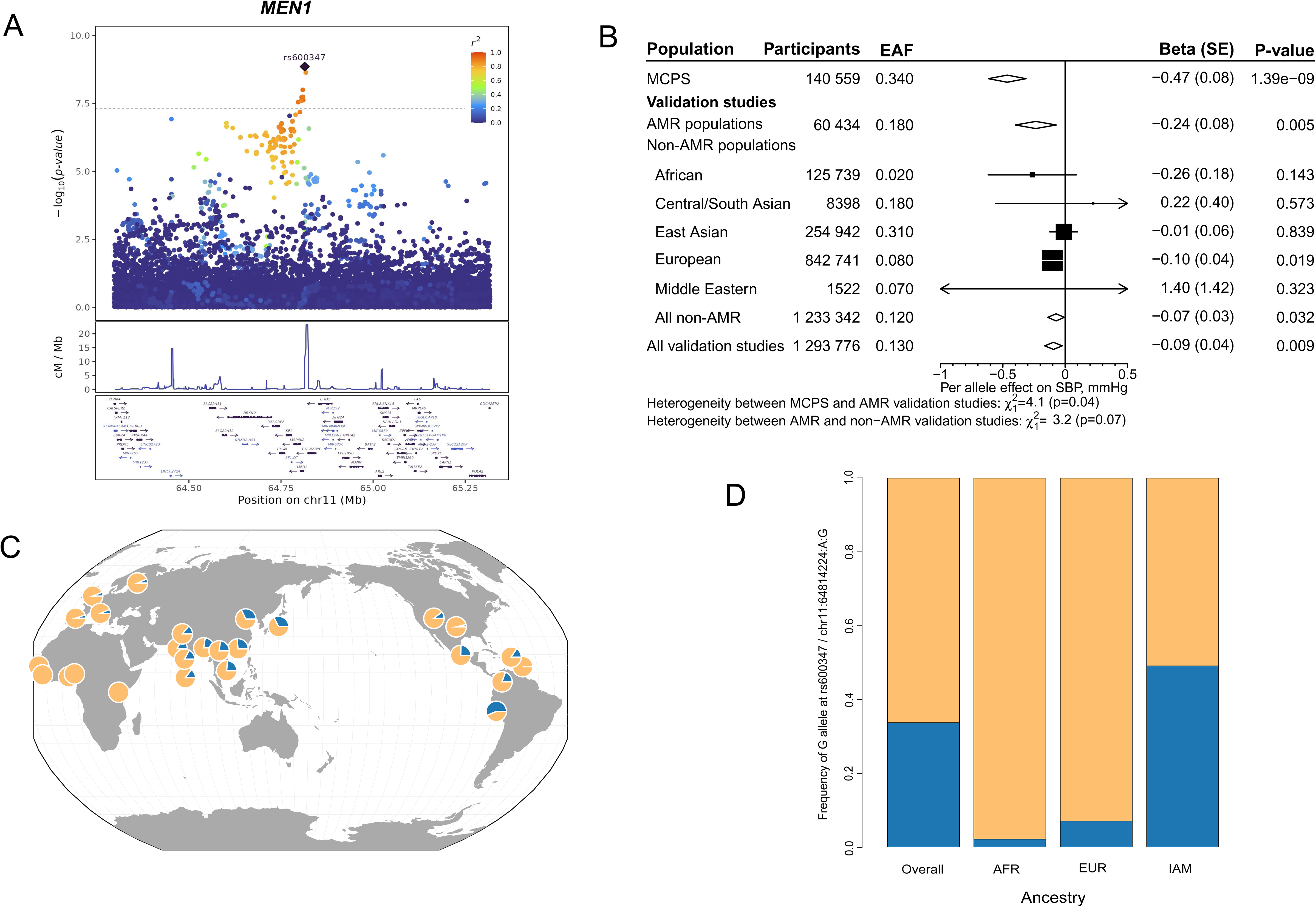
Effect of chr11:64814224:G:A / rs600347 on SBP. **A**: Locus plot for the novel SBP signal at *MEN1.* The y-axis shows −log_10_(*p)* values from approximate conditional analyses. Recombination rate and GENCODE gene annotations are shown below, with protein-coding and non-coding genes displayed in dark purple and blue, respectively. **B**: Meta-analysis of association of chr11:64814224:G:A / rs600347 with SBP in external cohorts. **C**: Relative frequencies of G (blue) and A alleles (yellow) at chr11:64814224:G:A / rs600347 in 1000 Genomes populations. Panel C generated with Geography of Genetic Variants Browser^51^. **D**: Relative frequencies of G alleles (blue) at chr11:64814224:A:G / rs600347 overall in MCPS and in genetic segments of AFR, EUR and IAM ancestry. AFR: African, AMR: admixed American, chr: chromosome, cM: centi-Morgan, EAF: effect allele frequency, EUR: European, IAM: Indigenous American, Mb: megabase, MCPS: Mexico City Prospective Study, SBP: systolic blood pressure, SE: standard error.

**Table 1:**
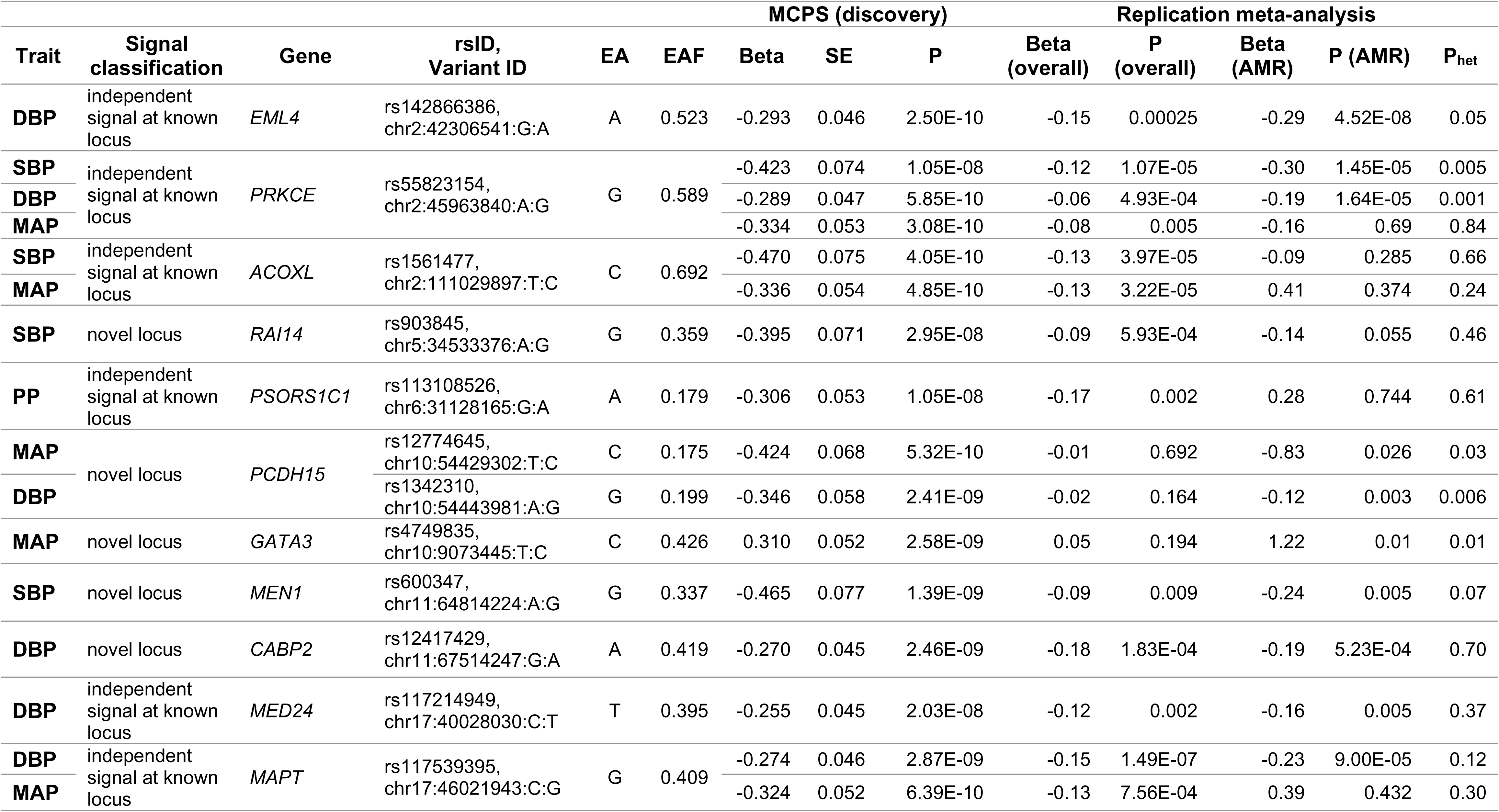
Novel signals replicating in either the overall, or AMR-ancestry meta-analysis of external cohorts. AMR: admixed American, DBP: diastolic blood pressure, EA: effect allele, MAP: mean arterial pressure, MCPS: Mexico City prospective study, P_het:_ heterogeneity p-value between overall and AMR-ancestry meta-analyses, PP: pulse pressure, rsID: reference single nucleotide cluster identifier, SBP: systolic blood pressure, SE: standard error.

Additionally, three further signals replicated only in AMR-ancestry cohorts. These were rs12774645 in an intron of *PCDH15* (effect size -0.18 mmHg vs -0.03 mmHg, p_het_=0.030, Supplementary Fig. 4), rs11213600 at a known locus near *CNTN5* (effect size -0.22 mmHg vs -0.02 mmHg, p_het_=0.005, Supplementary Fig. 5) and rs4072746 downstream of *NPTX1* (effect size 0.18 mmHg vs 0.02 mmHg, p_het_=0.039, Supplementary Fig. 6). All three have effect allele frequencies (EAFs) that differ between genetic segments of IAM and EUR ancestry, but rs12774645 is more common in EUR ancestry segments (Supplementary Table 16).

For DBP, MAP and PP there were one, five and two lead variants for potentially-novel signals not present in any replication dataset, respectively. These were largely rare variants more common in IAM-ancestry genetic segments (Supplementary Tables 18-20). Two variants associated with MAP (rs879737827 and rs193277537) were not rare in MCPS (with EAF of 0.011 and 0.069), but are roughly 40 and 20 times rarer in other ancestries on the gnomAD database^30^ (MAF 0.0022 in AMR vs. 0.00006 in EUR samples for rs879737827 and MAF 0.0392 in AMR vs. 0.0070 in EUR samples for rs879737827, respectively). In replication meta-analyses, 5 of the 11 remaining DBP signals (Table 1, Supplementary Fig. 7-11, Supplementary Table 21), 3 of 17 remaining MAP signals (Table 1, Supplementary Fig. 12-14, Supplementary Table 22), and one of three remaining PP signals replicated (Table 1, Supplementary Fig. 15, Supplementary Table 23). It should be noted that MVP PheWeb^22^, which provided the bulk of the AMR-ancestry sample size for replication, did not perform GWAS of MAP and PP, limiting power to replicate associations for variants that are rare in other ancestries.

Replicated signals included rs12417429 at a novel locus for DBP (Supplementary Fig.12), rs55823154 for both DBP and MAP (Supplementary Fig. 8 and 12) and rs113108526 for PP (Supplementary Fig.15). Two replicated DBP signals (rs142866386 and rs55823154) showed significant heterogeneity between AMR and non-AMR-ancestry meta-analyses (p_het_=0.0005 and p_het_=0.001, Supplementary Fig.7 and 8).

One further DBP signal (rs1342310) replicated only in AMR-ancestry meta-analysis (effect size -0.12 mmHg vs -0.003 mmHg for AMR-ancestry and non-AMR-ancestry meta-analyses respectively, p_het_=0.006, Supplementary Fig.16). Two further MAP signals also replicated only in AMR-ancestry; rs4749835 (effect size 1.22 mmHg vs 0.04 mmHg, p_het_=0.013, Supplementary Fig.17) and rs12774645 (effect size -0.83 mmHg vs -0.002 mmHg, p_het_=0.030, Supplementary Fig.18).

### Functional Integration

The processes through which genetic variants may influence BP traits were inferred in two ways. First, we used GARFIELD^31^ and chromatin accessibility annotations from the Common Metabolic Disease Genome Atlas^32^ to infer which overall patterns of tissues are more likely to harbour GWAS-significant variants in accessible chromatin. For SBP, the most significantly enriched annotation was accessible chromatin (assay for transposase-accessible chromatin using sequencing [ATAC-seq]) in the thoracic aorta (OR 6.33, 95% CI 3.94-10.2, Benjamini-Hochberg-adjusted p [p_BH_]=3.05×10^-11^). Other enriched tissues included kidney epithelial cells (OR 5.19, 95% CI 2.97-9.06, p_BH_=1.37×10^-7^), the adrenal gland (OR 6.63, 95% CI 3.03-14.5, p_BH_=7.56×10^-6^), and blood vessel endothelial cells (OR 4.48, 95% CI 2.90-6.93, p_BH_=2.87×10^-9^) and smooth muscle cells (OR 6.10, 95% CI 3.48-10.7, p_BH_=2.01×10^-8^, Supplementary Fig.

19).

GARFIELD analyses of DBP, MAP and PP identified similar enrichment patterns for tissues and cell types known to regulate blood pressure (Supplementary Fig. 19). For DBP, significant associations included adrenal cortex (OR 4.29, 95% CI 2.71-6.79, p_BH_=2.93×10^-7^), kidney (OR 5.39, 95% CI 2.81-10.2, p_BH_=6.37×10^-6^) and vascular smooth muscle (OR 5.26, 95% CI 2.83-9.78, p_BH_=3.83×10^-6^). Highly enriched tissues for MAP included the aorta (OR 5.00, 95%,CI 2.71-9.21, p_BH_=1.51×10^-6^), renal tract (OR 4.14, 95% CI 2.79-6.16, p_BH_=6.18×10^-10^) and adrenal gland (OR 4.33, 95% CI 2.87-6.54, p_BH_=6.70×10^-10^), and for PP included coronary artery (OR 5.56, 95% CI 2.73-11.3, p_BH_=6.01×10^-4^) and renal tract (OR 5.93, 95% CI 2.92-12.0, p_BH_=4.54×10^-4^).

Second, we assessed which tissues are most likely implicated at each locus using TACTICAL^33^, which integrates multiple annotations and the results of Bayesian fine-mapping. Assessment was run on chromatin state maps from 223 adult EpiMap biosamples^34^. TACTICAL analyses recapitulated known blood-pressure regulatory tissues (Fig. 4 and Supplementary Tables 24-27). For example, a signal for both SBP and MAP at the known *PDE3A* locus had the highest tissue of action score in the heart (0.79 for SBP and 0.76 for MAP). Further, a previously-reported signal for SBP at the *TBX3* locus had the highest tissue of action score in the adrenal gland (0.47). Third, a previously-reported SBP signal at the *PIK3CG* locus had the highest tissue of action score in the kidney (0.33).

**Figure 4:**
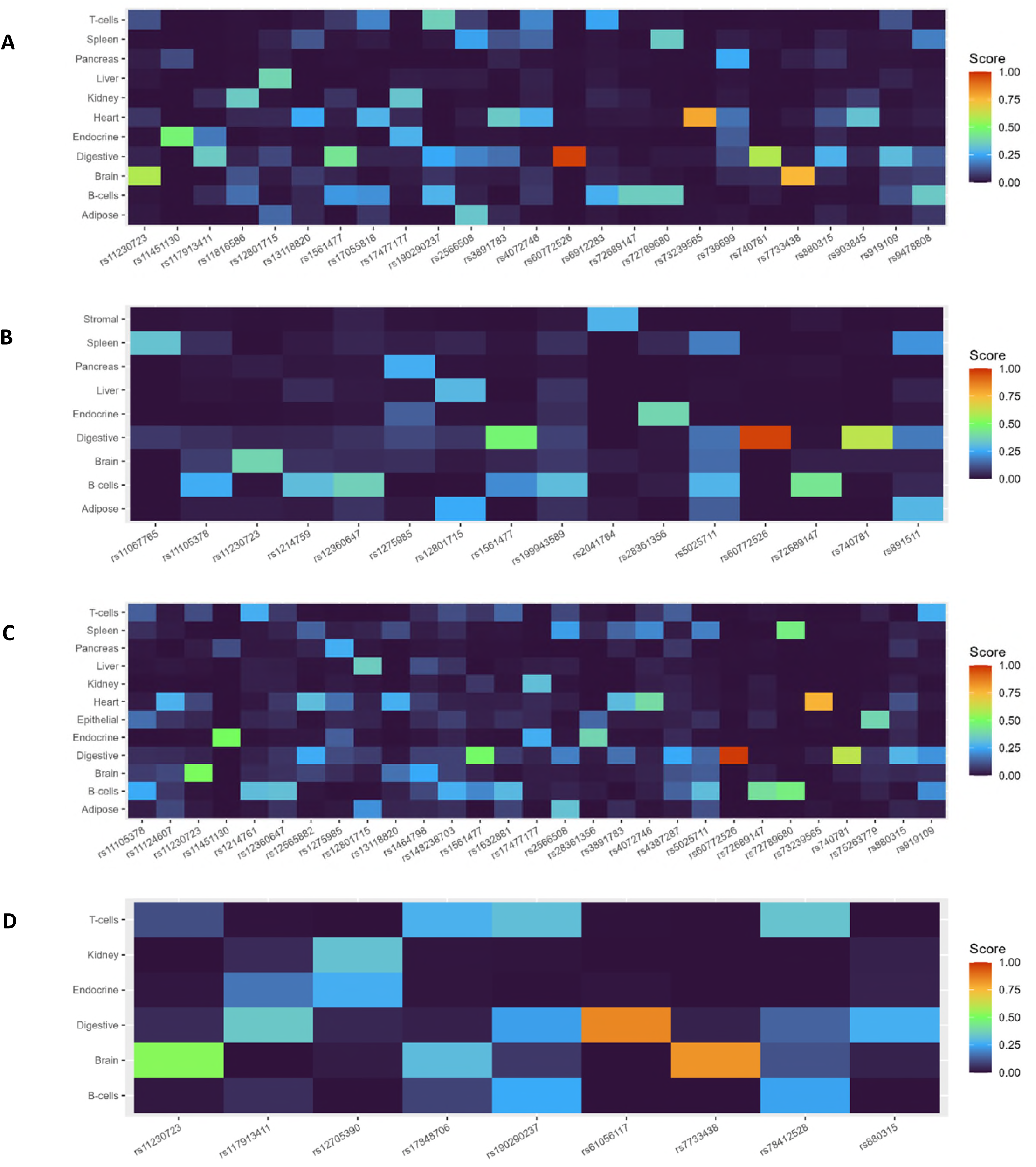
Tissue-of-action profiles for fine-mapped GWAS signals for systolic blood pressure. (A), diastolic blood pressure (B), mean arterial pressure (C) and pulse pressure (D) in MCPS, based on active chromatin states in 223 adult biosamples from EpiMap. TOA scores reflect the tissue specificity of each signal, based on the weighted partitioning of posterior probabilities from credible variants mapped to functional annotations. Results are collapsed into tissue groups and limited to tissues where ≥1 signal had TOA ≥ 0.25, and to signals with TOA ≥ 0.25 in at least one tissue.

Two strong TOA scores may have been impacted by using only adult annotation data. First, the SBP, MAP and PP signal at the *MEF2C* locus had the highest TOA score for brain. *MEF2C* plays a key role in brain development^35^, but also cardiovascular development^36^. Second, the signal at *HOXA13* associated with all four traits had highest TOA for the digestive tract. *HOXA13* is actively expressed in the adult digestive tract and likely plays a role in Barrett’s oesophagus^37^, but damaging variants can also lead a syndrome which includes urinary tract abnormalities, such as vesicoureteric reflux^38^ which may lead to kidney disease and elevated BP.

### Impact of rare coding variants on blood pressure

To explore the impact of coding genetic variation across the whole frequency spectrum on blood pressure traits, we leveraged exome sequencing (ES) data available in MCPS^18^ in two ways. First, we conducted exome-wide association studies (ExWAS) in REGENIE^39^ using 140,191 individuals with valid blood pressure measurements and ES data. ExWAS adjusted for the same set of variables as in the GWAS studies identified variants in numerous genes close to lead variants for GWAS signals (Supplementary Fig. 20). To identify rare, potentially-deleterious protein-coding variants not identified in GWAS, analyses were then restricted to rare variants (MAF<1%) and conditioned on lead hits for GWAS signals (Extended Data Fig. 2). For SBP and MAP, we identified a rare missense variant in *ZNF277* (chr7:112296259:A:G, rs747022720, MAF 0.013% in IAM-ancestry local ancestry segments, not observed in African or European-ancestry segments) that is predicted to cause a damaging aspartate to glycine substitution (SIFT score: 0.01 and PolyPhen score: 0.996, Table 2). Variants in *ZNF277* have previously been associated with kidney function^40^, a key physiological determinant of BP. In MCPS, the estimated glomerular filtration rate (eGFR) in carriers was 4.5 mL/min/1.73m^2^ lower than in non-carriers. However, only 27 alternative alleles were observed, and the difference was not statistically significant (p=0.26).

**Table 2:**
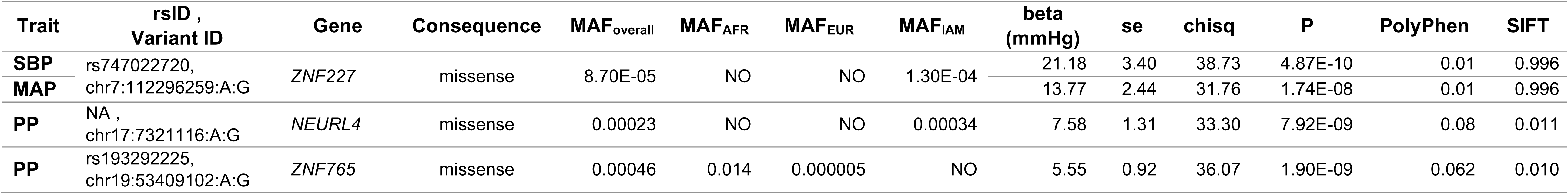
Significant associations for rare (MAF < 0.1%) variants with blood pressure traits in exome-wide association studies adjusted for lead GWAS hits. Displayed are variant details, canonical gene name, predicted consequence of the variant, overall and ancestry-specific MAF, association results and predicted tolerance by both PolyPhen (where values close to 0 imply strongly deleterious effects) and SIFT where values close to 1 imply strongly deleterious effects). AFR: African, EUR: European, GWAS: genome-wide association study, IAM: indigenous American, MAF: minor allele frequency, NA: not assigned, NO: not observed, rsID: reference single nucleotide cluster identifier, SE: standard error, SIFT: sorting intolerant from tolerant.

For PP, two rare variant associations were found. First, we identified a rare missense variant in *ZNF765* causing a damaging tyrosine to cysteine substitution (chr19:53409102:A:G, rs193292225, MAF 0.046%, SIFT score 0.01, Table 2). *ZNF765* variants have been found to be associated with measures of vascular endothelial function^41^. Second, there was a rare variant in *NEURL4* predicted to cause a likely tolerated leucine to proline amino acid substitution (chr17:7321116:A:G, no rsID assigned, MAF 0.034% in IAM-ancestry genetic segments, not observed in AFR or EUR-ancestry segments, SIFT score 0.08, PolyPhen score 0.011). Variants in *NEURL4* have been associated with SBP^22^.

Following ExWAS analyses, the impact of predicted loss of function, missense/protein altering variants with MAF <0.1% were analysed through gene-level tests across 18 153 protein-coding genes. To account for variants having a range of plausible effects on blood pressure traits, REGENIE’s Gene P test was used^42^. After correction for multiple testing, these analyses identified variants in *CCNI* that had large impacts on SBP (Supplementary Table 28). *CCNI* encodes cyclin-I, a protein essential to the cell cycle. The nine variants driving the association are rare frameshift variants predicted to have strong deleterious impacts on cyclin-I function. Mean SBP among 19 carriers of these variants was 141.3 mmHg vs 127.7 mmHg in non-carriers (Extended Data Fig. 4).

## Discussion

This work in 140,000 MCPS participants includes over twice as many AMR-ancestry participants as found in MVP^22^ and over three times as in All of US^23^. The study significantly enhances Latin American representation in GWAS of BP traits. The high proportions of Mesoamerican ancestry in MCPS allows examination of genetic variation not possible elsewhere^17,18^.

We identified 113 distinct independent signals, with 78, 61, 88 and 24 associated with SBP, DBP, MAP and PP, respectively (Fig. 2, Supplementary Table 10). The strongest associations corroborated those observed in other populations, such as signals at *FGF5* and *KCNK3*. In total, 60 signals either identified novel loci or were conditionally-independent of all previously reported associations with any BP trait. The findings add to the total of 2103 signals identified in over 1 million individuals of European ancestry^9^. Variants at novel signals in our work were overwhelmingly more common in IAM-ancestry genetic segments. For example, the novel variant at rs600347 has an EAF of 0.49 in IAM-ancestry segments, but 0.02 and 0.07 in EUR and AFR ancestry-segments, respectively (Supplementary Table 16 and 18-20), and 13 replicated in external cohorts (Table 1).

Nine novel signals showed significant heterogeneity between AMR and other ancestries in replication meta-analyses (Fig. 2A, Supplementary Fig. 4-8 and Supplementary Fig. 16-18). Previously-reported variants showed no significant heterogeneity between MCPS and GWAS in other ancestries (Supplementary Fig. 3), suggesting that differences in LD may be a key factor. Our findings demonstrate that differences in allele frequencies and LD allow novel discoveries in under-represented ancestries with sample sizes far smaller than used in the largest European-ancestry BP GWAS^9^.

Among the novel findings was a signal for SBP with lead variant rs600347, located upstream of tumour suppressor gene *MEN1*. Inactivating variants in *MEN1* cause the Mendelian condition, multiple endocrine neoplasia type 1 (MEN1). This disorder can produce pituitary adenomas secreting adrenocorticotropic hormone (ACTH) and adrenocortical tumours secreting cortisol or aldosterone. Each of these hormones can raise blood pressure^43^. The GTEx catalog lists *MEN1* eQTLs in the aorta and adrenal gland for this variant. Additionally, variants at *MEN1* have been associated with serum urate levels^44^ which are influenced by traits such as adiposity and kidney function that may also influence BP. The greater prevalence of the variant in AMR-ancestry individuals made it much easier to detect that in other ancestries.

The DBP signal with lead variant rs12417429 lies downstream of *CABP2*, a calcium-binding protein expressed in the heart and arterial tree. Finally, also notable is the MAP signal near *GATA3*, a transcription factor with important roles in endothelial cell function^45^ and kidney development^46^.

Novel signals with plausible roles in blood pressure are especially interesting because they may lead to new biological understanding and therapeutic targets. Advances could benefit not just people of AMR ancestry; discovery of PCSK9 inhibitors in people of African ancestry led to cholesterol-lowering drugs benefiting people of all ancestries^47,48^.

Functional integration analyses supported the importance of known blood pressure-regulating pathways. Both GARFIELD and TACTICAL found the heart and arteries to be key regulatory tissues. GARFIELD additionally identified the adrenal gland and kidney as important regulatory organs, in keeping with the known physiology of blood pressure^2^. TACTICAL analyses also identified signals at *MEF2C* and *HOXA13* to have highest tissue-of-action scores in the brain and digestive tract, respectively. In both these examples, varied expression of transcription factors between embryonic development and adulthood mean that greater availability of embryonic biosamples might allow more precise delineation of the tissues of action.

Rare variant ExWAS analyses conditioned on GWAS results identified rare missense variants in *ZNF277* associated with SBP, and in *ZNF765* and *NEURL4* associated with PP. *ZNF277* and *ZNF765* encode zinc finger proteins, which influence gene expression. *NEURL4* encodes a centriole protein thought to be important in proteasomal degradation within the cell. Variants in these genes have been associated with traits that may influence BP (kidney function for *ZNF277* and vascular endothelial function for *ZNF765*) or a BP trait itself (SBP for *NEURL4*).

Gene-based tests identified predicted loss of function (pLoF) variants in *CCNI* as influencing blood pressure. *CCNI* is highly conserved and encodes cyclin-I, a cell cycle protein. *CCNI* variants identified here have been associated with height^49^, but not with other traits. A biological mechanism for the blood pressure relationship is not immediately clear.

A limitation of our study was that blood pressure measurements (though multiple) were obtained at a single point in time. Previous work has shown that repeat measures allowing analysis of usual blood pressure accounts for substantial intra-individual variability and doubles the variance in blood pressure traits explained by genetic variants^50^. Second, functional integration analyses are limited by a dearth of AMR-ancestry datasets. Inclusion of more diverse ancestries in eQTL and pQTL datasets will enhance the breadth of populations in which well-powered functional integration analyses can be conducted. Third, replication of potentially-novel blood pressure associations would benefit from larger AMR-ancestry GWAS datasets.

Our study demonstrates that genetic analyses in Latin American populations may lead to novel discoveries and corroborate findings made in other populations. Publicly-available results generated by this study will further open science initiatives to alleviate the burden of BP-related diseases in Latin America and its diaspora.

## URLs

bigsnpr (R package) https://cran.r-project.org/web/packages/bigsnpr/

CMDGA https://cmdga.org/

dbSNP https://www.ncbi.nlm.nih.gov/snp/

GARFIELD https://www.ebi.ac.uk/birney-srv/GARFIELD/

MCPS http://www.ctsu.ox.ac.uk/research/mcps

NHGRI-EBI GWAS Catalog https://www.ebi.ac.uk/gwas/

refTSS annotations: http://reftss.clst.riken.jp/

SUGEN https://github.com/dragontaoran/SUGEN/

TACTICAL https://github.com/Jmtorres138/TACTICAL

TOPMed Imputation Server https://imputation.biodatacatalyst.nhlbi.nih.gov/

## Online Methods

### Recruitment, follow-up and genotyping

The Mexico City Prospective Study (MCPS) was established by scientists at the National Autonomous University of Mexico (Universidad Nacional Autónoma de México; UNAM) in collaboration with scientists at the University of Oxford with the aim of identifying major risk factors for disease and premature death in Mexico. As described previously, MCPS^52^ recruited 159 755 adults aged 35 years or older via household survey in the Coyoacán and Iztapalapa districts of Mexico City from 1998-2004. Demographic, health and lifestyle data, and physical measurements including height, weight and three sitting blood pressures at one-minute intervals were collected by trained survey staff.

Participants gave a 10mL non-fasting blood sample in an ethylene-diamine-tetra-acetic acid (EDTA) tube which was centrifuged to yield plasma and buffy coat layers. DNA was extracted and sent to the Regeneron Genetics Center (RGC; New York, USA) for genotyping of 650,380 variants on the second-generation Illumina Global Screening Array (GSAv2; Illumina, California, USA).

Genotyping yielded data on 650,380 variants in 140,831 individuals after quality control (QC), as described previously^18^. Imputation using the Trans-Omics Precision Medicine (TOPMed) version R2 in the GRCh38 imputation server^53^ yielded data on 42,121,162 variants with imputation r^2^ scores ≥0.4 and N_eff_≥30.

### Phenotype selection

Participants with missing or extreme blood pressure data (SBP <80MMHG or >250mmHg, DBP <40MMHG or >150mmHg, PP <15mmHg)^54^ or DNA not passing QC were excluded. Participants taking an antihypertensive medication had their SBP and DBP increased by 15mmHg and 10mmHg, respectively, as is standard in the field^55,56^.

### Principal component analysis

Principal components (PCs) were generated from genotyped autosomal variants passing QC using the *bigsnpr* workflow^57^ using a clumping r^2^ threshold of 0.005 and a minor allele frequency (MAF) threshold of 5%. PCs were calculated in an unrelated set of 40,695 individuals and the remaining MCPS participants projected onto the PCs. Examination of loading plots demonstrated that the first seven PCs incorporated variants from across the genome and described meaningful structure. These were used to adjust for population structure.

### GWAS and estimating genomic inflation

Population effect sizes for variants passing QC were estimated using SUGEN^24^ adjusting for age, age-squared, biological sex, body mass index (BMI), and the first seven genetic principal components^54^. SUGEN uses generalised estimation equations to estimate effect sizes first within pedigrees then across them, thereby adjusting for complex patterns of relatedness. It was designed for use in related samples of admixed populations, such as MCPS. First degree pedigrees were inferred from PRIMUS,^58^ and SUGEN was run without sampling adjustments as no over-sampling was incorporated into the recruitment strategy. The genome-wide significance threshold was set at p<5×10^-8^ and a locus was defined as a 1 megabase (Mb) region of the genome; i.e. 500 kilobases (kb) either side of the lead variant^59^.

Genomic inflation factors, λ_GC_, were calculated from the median test statistic. Covariate-adjusted linkage disequilibrium score regression^25^ (cov-LDSC) was as used to estimate genomic inflation with LD scores generated from 9764 whole-genome sequenced (WGS) MCPS participants^18^ unrelated at a KING kinship threshold of 0.0442. PCs were generated for this set of MCPS participants as described above for (but without the need for projection). Cov-LD scores were calculated for 18,789,983 variants with MAF≥0.1% (in keeping with the threshold used in the original LDSC paper^60^) in windows of 1Mb (in keeping with the window size used in the pan-UK biobank^61^).

### Conditional analyses

Sets of conditionally-independent variants were identified using conditional joint analysis implemented in the Genome-wide Complex Trait Analysis software package (GCTA-COJO)^62,63^. Loci associated with blood pressure traits were delineated by defining a 500kb region around each significantly-associated variant. Where these regions overlapped, they were combined into a single region. Within each locus a set of conditionally-independent variants was then identified through forwards-selection-backwards-elimination using GCTA as described by Mahajan *et al*^64^.

First, forward-selection was carried out. Variants at each locus were conditioned on the lead variant at each locus and retained if conditionally-independent until no further significant variants were found. For backwards-elimination, the set of significant variants was included in a joint analysis and those with joint p-values not reaching the 5×10^-8^ significance threshold removed. Forward-selection and backwards-elimination were repeated until a stable set of variants was identified. Variants identified as conditionally-independent of each other in this way were considered independent signals within a locus.

### Fine mapping

For each signal selected through forward-selection-backwards-elimination, Bayesian fine-mapping was carried as per Mahajan *et al*.’s^64^ implementation of Wakefield’s Bayes’ false-discovery probability methodology^65^. Variants were restricted to those with MAF≥0.25%. The approximate Bayes’ factor was calculated as:

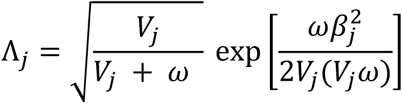

Where β_j_ is the estimated effect of variant j, V_j_ is the variance of the effect estimate and ω is the prior variance (set at 0.04 as per Wakefield^65^).

The posterior probability of variant j (π_j_) (of k analysed variants) driving the association was given by:

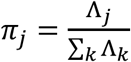

The variant with the highest π_j_ for each signal was designated the lead variant. 99%-credible sets were calculated by ordering variants by descending posterior probability and including variants until the cumulative posterior probability reached 0.99.

### Heterogeneity by ancestry

For variants previously reported as associated with any blood pressure trait in the GWAS Catalog^66^, associations from the study with the largest sample size not containing Latin American-ancestry individuals, and reporting a 95% confidence interval, was selected and compared to associations from MCPS.

Z scores for differences in effect sizes between MCPS and GWAS Catalog associations were computed as per Mahajan *et al*.^64^:

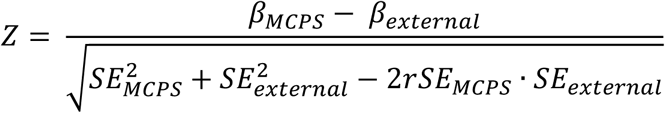

Where β_MCPS_ and β_external_ are the effect sizes from the MCPS and GWAS Catalog analyses respectively, SE is the standard error of the respective estimates and *r* is the estimated correlation between β_MCPS_ and β_external_.

### Gene annotations

For each signal, the lead variant was annotated with the Ensembl Variant Effect Predictor (VEP) and Ensembl version 113 for genome build GRCh38^67^. This identified the nearest canonical gene, and, in the case of missense coding variants, the sorting intolerant from tolerant (SIFT)^68^ score. Allele frequencies for the effect allele were calculated for the overall cohort from the maximum unrelated set of 79,612 MCPS participants, and for the three major continental ancestries present in MCPS (West African [AFR], European [EUR]and Indigenous American [IAM]) from local ancestry segments inferred from RFMix as described previously^18^, and as listed on the MCPS variant browser (**URLs**).

### Novel associations

Previously-reported associations with any of the four blood pressure traits or hypertension were identified in the GWAS Catalog^66^. Associations were designated as a potentially novel loci if they were (1) >500kb from any previously-reported associations, and (2) had at least two supportive locus-wide significant associations (p<5×10^-5^)^69,70^ within 500kb in the MCPS GWAS results. Variants were deemed indeterminate if they were >500kb from known associations but had no supportive variants. Known loci were designated as those within 500kb of a previously-reported association.

Potentially novel associations independent of all reported associations at known loci were sought via conditional analyses. PLINK2.0^71^ was used to calculate LD r^2^ between previously-reported variants and MCPS signals in a maximum unrelated set of 79,612 MCPS participants unrelated at the 3^rd^ degree or closer. If a variant was not present in MCPS, then the LD r^2^ was taken from LDpair using GRCh38 High Coverage in All Populations^72^. Conditionally-independent variants with no reported variants in LD at r^2^≥0.9 in MCPS were taken forwards for conditional analyses. These were run in GCTA v1.9^62^ using the --cojo-cond and --cojo-colinear 0.9 options. Signals attaining a conditional p-value <5×10^-8^ were considered potentially-novel associations at known loci.

### Replication

Replication was sought in publicly-available results from China-Kadoorie Biobank (CKB)^73^, Million Veteran Program (MVP)^22^, Population Architecture using Genomics and Epidemiology (PAGE)^12^, and pan-UK Biobank (pan-UKB)^74^ studies. However, none of the novel variants were present in PAGE. Further replication was sought by directly approaching the Multi-Ethnic Study of Atherosclerosis (MESA)^75^.

MESA is a study of the characteristics of subclinical cardiovascular disease and the risk factors that predict progression to clinically overt cardiovascular disease or progression of the subclinical ^75^. MESA consists of a diverse, community-based sample of an initial 6,814 men and women aged 45-84 years without known cardiovascular disease at baseline. Thirty-eight percent of the recruited participants were White, 28 percent African American, 22 percent Hispanic, and 12 percent of Chinese descent. Participants were recruited from six field centres across the United States: Baltimore City and Baltimore County, Maryland; Chicago, Illinois; Forsyth County, North Carolina; Los Angeles County, California; New York, New York; and St. Paul, Minnesota. The first examination took place over two years, from July 2000 to July 2002, and has been followed by additional examinations.

Participants are being followed for identification and characterization of cardiovascular disease events, including acute myocardial infarction and other forms of coronary heart disease (CHD), stroke, and heart failure; for cardiovascular disease interventions; and for mortality. Follow-up telephone interviews with participants or their proxies are attempted at least annually to identify new hospitalizations and diagnoses. MESA staff request information on hospital admissions for any reason, outpatient CVD diagnoses, and death. Death certificate records are obtained from state vital statistics or the National Death Index. MESA staff record International Classification of Disease (ICD) diagnosis codes for all hospitalizations and request medical records for those with ICD codes related to CVD. MESA staff also request outpatient records for potential CVD events. However, not all hospitalizations and/or CVD event records were fully documented.

The study was approved by the Institutional review boards at all participating institutions, and all participants gave written informed consent. In addition, informed consent was obtained for extensive data sharing (dbGaP) and genetic/omic studies, including candidate genes (NHLBI CARe), genome-wide scans (NHLBI SHARe), exome sequencing (NHLBI ESP) and, most recently, the NHLBI TOPMed program.

Inverse-variance-weighted meta-analysis of all replication cohorts was used to estimate genetic variant effects externally. False-discovery rate (FDR) adjustment was conducted using the Benjamini and Hochberg method^76^. Heterogeneity across replication datasets was assessed using Cochrane’s Q test^77^. Potentially-novel variants were considered to have replicated if they had a concordant direction of effect with the MCPS discovery GWAS and an FDR-adjusted p-value<0.05.

### Functional integration

Tissues in which GWAS-significant signals were likely to act were sought using two methods. First, GWAS Analysis of Regulatory or Functional Information Enrichment with LD correction (GARFIELD)^31^ was run to identify tissue-specific annotations more likely to harbour GWAS-significant results. GARFIELD takes a set of independent variants (LD r^2^ < 0.01) and annotates them (or proxies [LD r^2^ ≥ 0.8]) if they lie in a region with a given tissue-specific annotation. It then runs logistic regression models where the outcome is whether a given p-value threshold is met and independent variables are distance to the nearest transcription start site (TSS), MAF and number of LD proxies (LD r^2^ ≥ 0.8). This is run for each annotation in turn, and then a multi-annotation model is run to identify a set of independent annotations associated with variants reaching a p-value threshold.

Tag and minor allele frequency transcription start site distance (*maftssd*) files were generated from MCPS data and TSS data for humans in GRCh38^78^ from refTSS (**URLs**). Annotation information was generated from genomic annotation Bed files in GRCh38 hosted on the Common Metabolic Diseases Genome Atlas (CMDGA)^79–82^.

GARFIELD was run on variants with MAF ≥0.1% calculating single annotation enrichment at 5×10^-5^, 1×10^-6^ and 5×10^-8^ p-value thresholds. For the multi-annotation step, the 5×10^-8^ threshold was used with the default condition and adjusted p-value thresholds.

Second Tissue of Action scores for Investigation Complex trait-Association Loci (TACTICAL)^33^ was used to systematically integrate 99% genetic credible sets from fine-mapping analyses (see above) with molecular epigenomic maps. Chromatin state maps generated by the EpiMap Project^81^, corresponding to the 18-state Roadmap model, were obtained for 223 adult biosamples (28 unique tissues) from CMDGA. Chromatin states in this model were based on ChIP-seq data for six histone marks: H3K27ac, H3K4me1, H3K4me3, H3K36me3, H3K9me3, and H3K27me3. Ten states were used for calculating TOA scores and were ordered into three tiers based on the emission probability strength for H3K27ac. Tier 1 included strong enhancer and promoter elements (“TssA”, “TssFlnkU”, “EnhG2”, and “EnhA1”), and each state was assigned a weight of “3” in the TACTICAL analysis. Tier 2 included state “EnhA2” which was given a weight of “2”. Tier 3 included weak enhancer and promoter elements (“TssFlnk”, “TssFlnkD”, “EnhG1”, and “EnhWk”) and actively transcribed elements (“Tx”) and each state was given a weight of “1”. The cumulative PPA value for each fine-mapped GWAS signal was then partitioned into TOA scores using the specified annotation weights, and analyses were performed for both the level of whole tissues and individual biosamples or cell types. An “Unclassified” score corresponded to the cumulative PPA value of credible variants that did not map to any of the epigenomic annotations across tissue and cell types.

### Exome-wide association studies

As in previous work^83^, analyses of exome sequencing data from 141,045 MCPS participants were restricted to a set of 9,325,897 variants that were previously selected from a QC procedure that involved both machine-learning based detection of low-quality variants and hard filters (e.g. variant missingness >10%, HWE departure *p-value* <1×10^-30^, etc.)^18^. Single-variant and gene burden tests were performed using REGENIE v3.1.3. The set of quality-controlled genotyping array variants was further restricted to those with MAC ≥100 (m = 560,015 variants; n = 140,829 individuals) and used to fit the whole-genome regression model in step 1 with a block size of 1000. In step 2, a linear regression score test was implemented to test for single-variant associations with BP traits and incorporated the genome-wide predictors estimated in step 1 in a leave-one-chromosome-out (LOCO) procedure. For step 2, a block size of 400 was used and only variants with a minimum MAC ≥25 were tested. Association tests in step 2 were also performed conditional on all genome-wide significant variants present within the exome variant set with the --condition-list argument and restricted to variants with minor allele frequency <1% (which are more likely to have deleterious effects). All genetic association tests were adjusted for age, age-squared, sex, BMI, and the first seven genetic PCs. Significant variants were annotated using the Ensembl VEP web interface, which collated information on allele frequencies from the 1000 Genomes Project (1KG) and gnomAD exome and whole-genome sequencing data, variant consequences for MANE Select transcripts, and variant pathogenicity scores from SIFT, PolyPhen, PrimateAI, REVEL, and CADD.

### Gene burden tests

As previously^77^, exome sequence variants were annotated per guidelines from the Biobank Rare Variant Analysis (BRaVa) Consortium (https://github.com/BRaVa-genetics/variant-annotation), which integrates VEP predictions, CADD scores, and splice-site disruption scores from SpliceAI. Variant annotations were restricted to canonical, protein-coding transcripts and stratified into five mutually-exclusive categories: (1) <u>Putative loss-of-function</u> (pLoF) variants with a high-confidence loss-of-function (LoF) call as per Loftee (m = 181,675 variants); (2) <u>Damaging missense or protein altering</u> variants meeting any of the following thresholds; a) REVEL score ≥ 0.773; b) CADD_PHRED_ ≥ 28.1; c) SpliceAI max Δ-score ≥ 0.20; low-confidence LoF call as per Loftee (m = 520,998 variants); (3) <u>Other missense or protein-altering</u> variants, i.e. missense, start-loss, stop-loss, or in-frame indel variants not categorized in (2) (m = 2,118,452 variants); (4) <u>Synonymous</u> variants with SpliceAI DS score <0.2 (m = 1,195,125 variants); (5) <u>Non-coding</u> variants impacting UTRs, introns, or intergenic regions (m = 6,897,946 variants). Variants that were not classified in any of these five categories were omitted from further analyses.

Gene masks in the rare variant burden tests included either pLOF variants (category 1) or pLoF or damaging missense or protein-altering variants (categories 1 and 2). Two allele frequency filters were also applied: singletons or variants with MAF <0.1%, resulting in a total of four masks per gene. Gene-based tests for SBP, DBP, MAP and PP were performed as implemented in REGENIE v3.1.3 with the same genome-wide predictors and linear model covariates as used for the ExWAS. We performed the “Gene-P” combination test that utilises the results of four separate gene-based tests – SBAT, SKAT-O, ACAT-V, and BURDENT-ACAT – to evaluate gene-level associations, where each test applies a different strategy for grouping and weighting variants. The Cauchy method is then applied within the Gene-P test to combine results into a single p-value per gene. Genes with a Gene-P *p*-value <1.42×10^-6^ (i.e. Bonferroni p-value correcting for 17,576 protein-coding genes with results and two traits, as BP traits are correlated) were deemed to be significantly associated.

## Ethics

Ethics approval was granted by the National Council of Science and Technology in Mexico, the Mexican Ministry of Health, the Ethics and Research commission from the Medicine Faculty at the National Autonomous University of Mexico (UNAM) and the University of Oxford.

## Funding

MCPS has received funding from the Mexican Health Ministry, the National Council of Science and Technology for Mexico, Wellcome Trust (058299/Z/99), Cancer Research UK, British Heart Foundation (RE/13/1/30181), Kidney Research UK, and the UK Medical Research Council (MC_UU_00017/2, MR/Z504543/1).

MESA and the MESA SHARe project are conducted and supported by the National Heart, Lung, and Blood Institute (NHLBI) in collaboration with MESA investigators. Support for MESA is provided by contracts 75N92025D00022, 75N92020D00001, HHSN268201500003I, N01-HC-95159, 75N92025D00026, 75N92020D00005, N01-HC-95160, 75N92020D00002, N01-HC-95161, 75N92025D00024, 75N92020D00003, N01-HC-95162, 75N92025D00027, 75N92020D00006, N01-HC-95163, 75N92025D00025, 75N92020D00004, N01-HC-95164, 75N92025D00028, 75N92020D00007, N01-HC-95165, N01-HC-95166, N01-HC-95167, N01-HC-95168, N01-HC-95169, UL1-TR-000040, UL1-TR-001079, UL1-TR-001420, UL1TR001881, and R01HL105756. The authors thank the MESA participants and the MESA investigators and staff for their valuable contributions. A full list of participating MESA investigators and institutions can be found at http://www.mesa-nhlbi.org.

## Data availability

Data from the Mexico City Prospective Study are available to *bona fide* academic researchers. For more details, the study’s Data and Sample Sharing policy may be viewed (in English or Spanish) at https://www.ctsu.ox.ac.uk/research/mcps. Available study data can be examined in detail through the study’s Data Showcase, available at https://datashare.ndph.ox.ac.uk/mexico/. MCPS ancestry-specific allele frequencies are available in a public browser (https://rgc-mcps.regeneron.com/).

## Competing Interests

JRE declares cohort funding from Regeneron and AstraZeneca to the University of Oxford. RH declares trial funding from Boehringer Ingelheim to the University of Oxford. NS declares institutional grant funding from Boehringer Ingelheim, Lie Lily and Novo Nordisk.

---

**Extended Data Figure 1:**
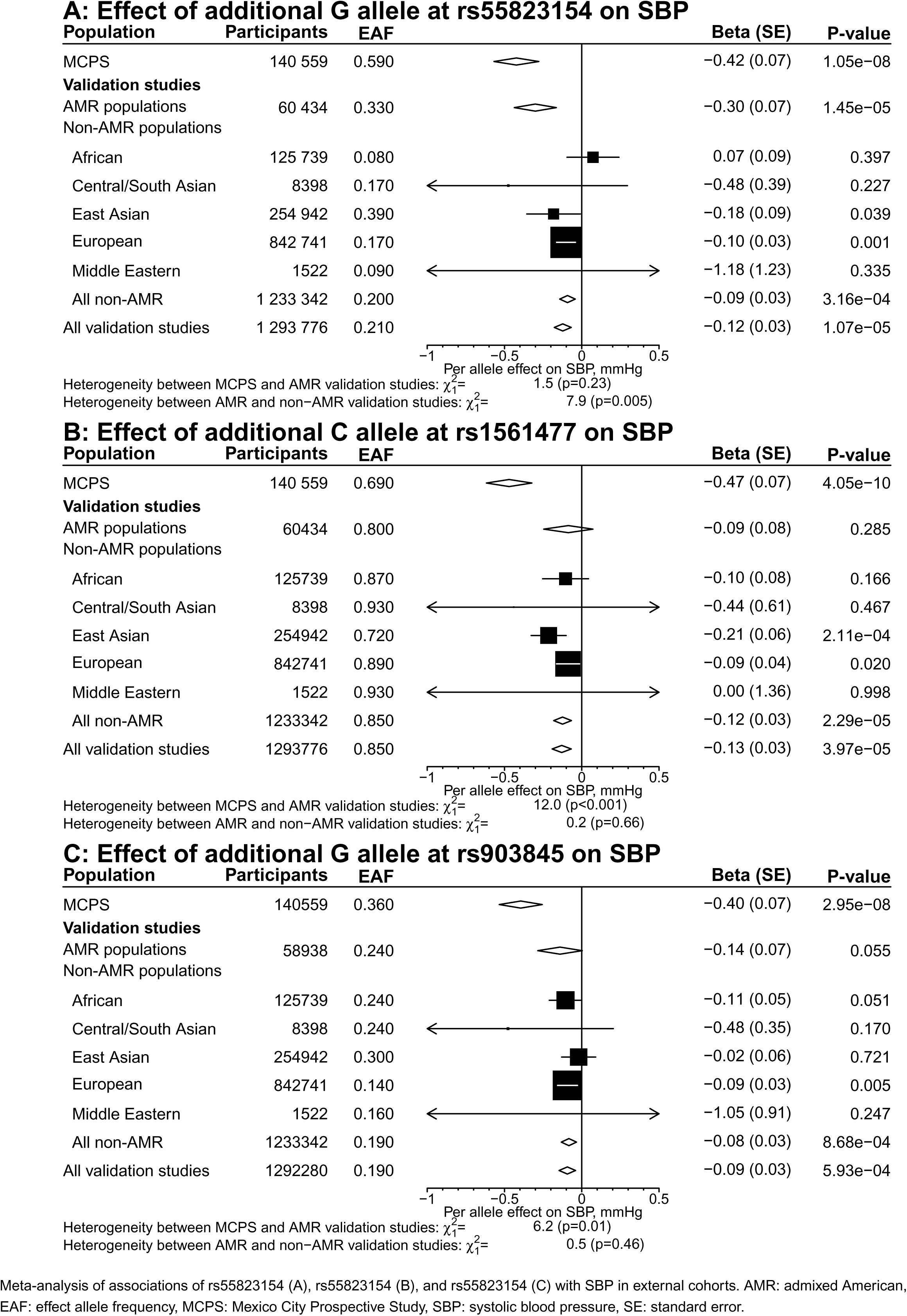
SBP signals replicating in external meta-analyses. Meta-analysis of associations of rs55823154 (A), rs903845 (B), and rs1561477 (C) with SBP in external cohorts. AMR: admixed American, EAF: effect allele frequency, MCPS: Mexico City Prospective Study, SBP: systolic blood pressure, SE: standard error.

**Extended Data Figure 2:**
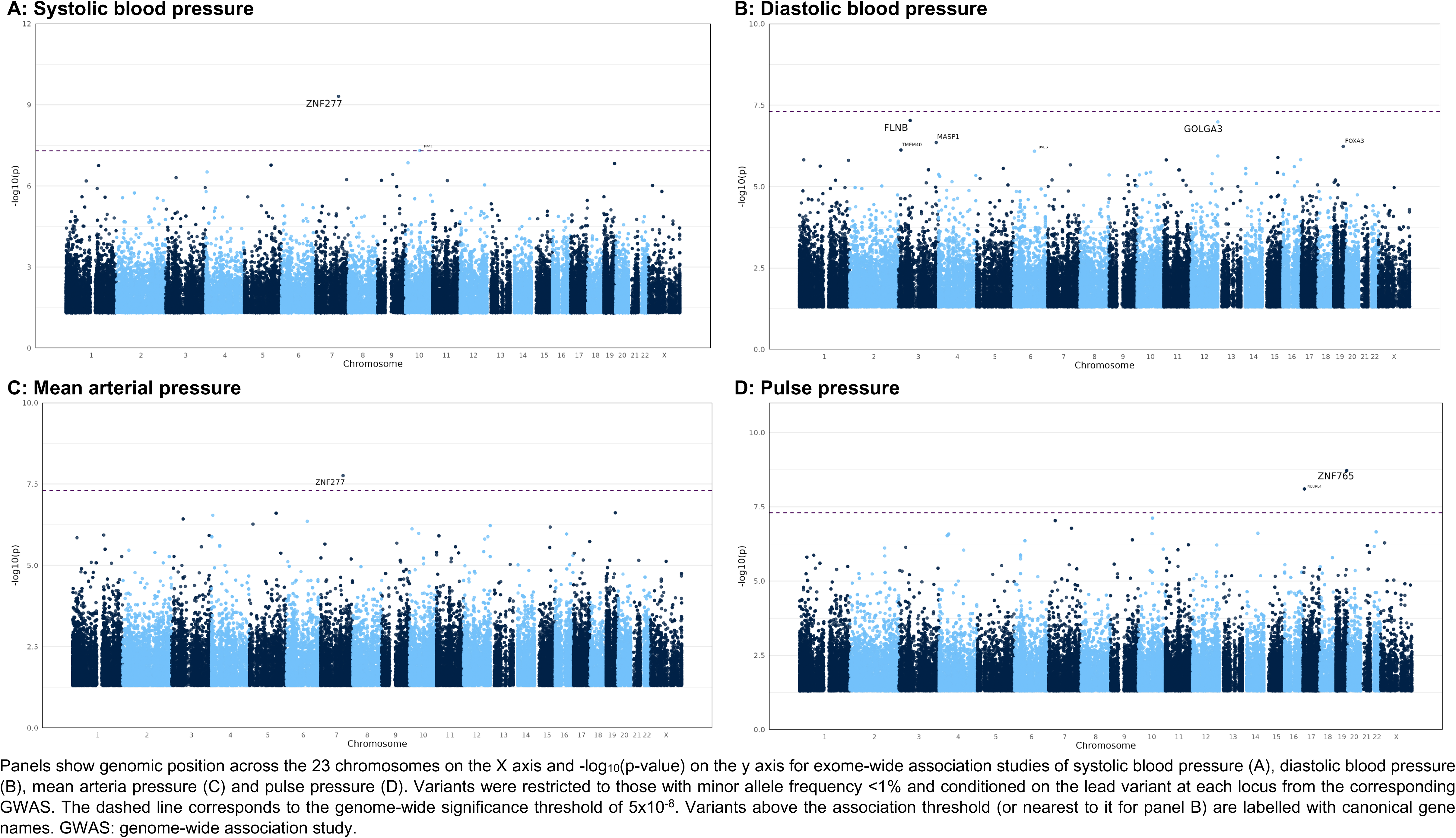
Manhattan plots of rare variant exome-wide association studies of blood pressure traits, conditioned on lead GWAS variants. Panels show genomic position across the 23 chromosomes on the X axis and -log10(p-value) on the y axis for exome-wide association studies of systolic blood pressure (A), diastolic blood pressure (B), mean arterial pressure (C) and pulse pressure (D). Variants were restricted to those with minor allele frequency <1% and conditioned on the lead variant at each locus from the corresponding GWAS. The dashed line corresponds to the genome-wide significance threshold of 5×10-8. Variants above the association threshold (or nearest to it for panel B) are labelled with canonical gene names. GWAS: genome-wide association study.

**Extended data Figure 3:**
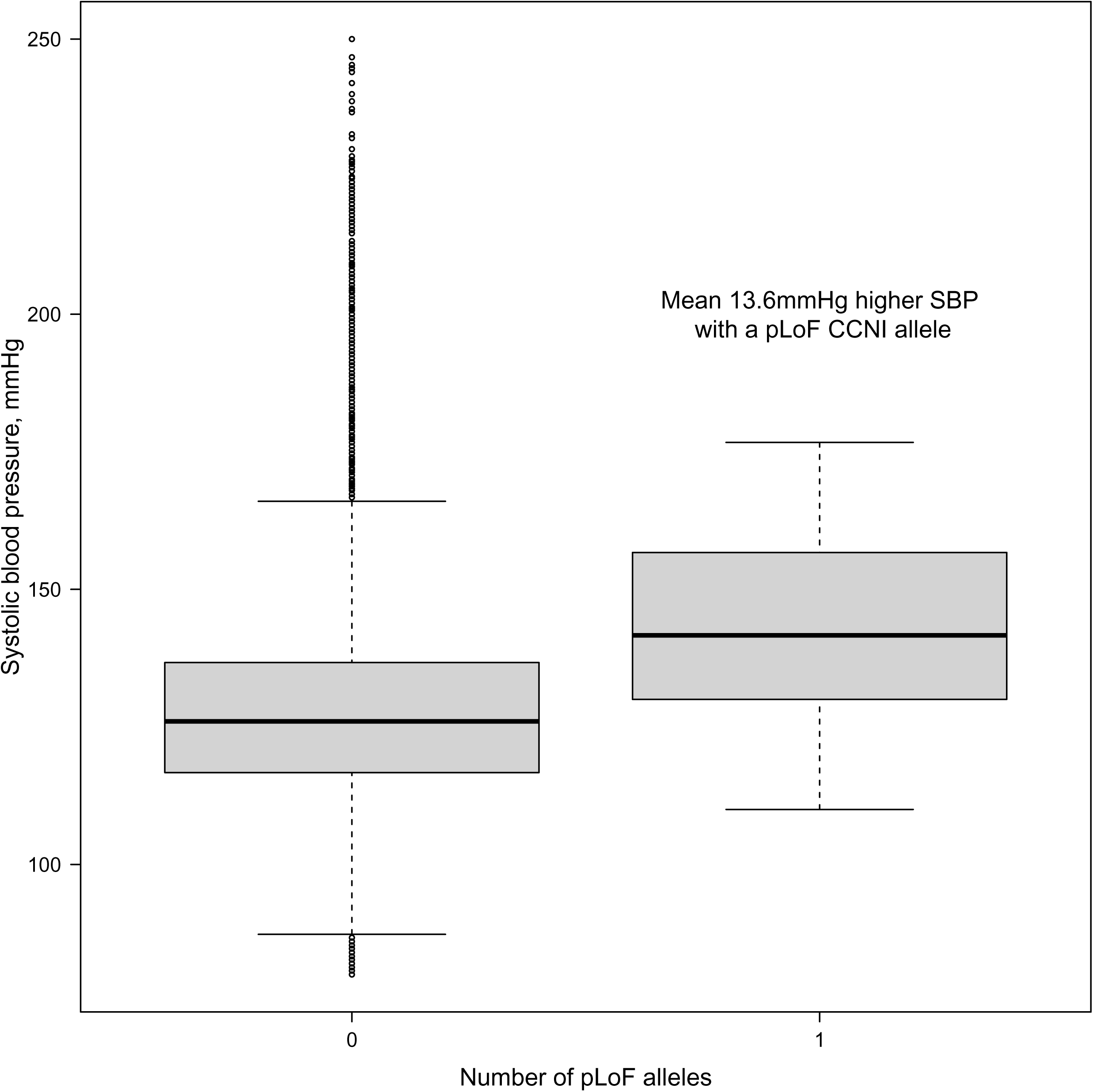
SBP by CCNI carrier status. SBP in MCPS participants without or with variants with a high−confidence pLoF call as per Loftee. MCPS: Mexico City Prospective Study, pLoF: putative loss−of−function, SBP: systolic blood pressure.

